# Early-life infectious disease exposure, the “hygiene hypothesis”, and lifespan: evidence from hookworm

**DOI:** 10.1101/2024.12.09.24318730

**Authors:** Ralph I. Lawton

## Abstract

Exposure to infectious disease in early life may have long-term ramifications for health and lifespan. However, reducing pathogen exposure may not be uniformly beneficial. The rise of modern sanitation and reduction of infectious diseases has been implicated in increasing levels of allergy and immune dysregulation: termed, the “hygiene hypothesis.” This study leverages quasi-experimental variation from combining pre-campaign hookworm exposure with the Rockefeller Sanitary Commission’s de-worming campaign in the early 20th century to rigorously examine the impacts of childhood hookworm exposure on adult lifespan and morbidity. Findings show de-worming before age five leads to 2.5 additional months of life in a large sample of adult death records. Further, decreasing hookworm exposure is related to improvements in biomarkers for inflammation and skin-tested allergies, in contrast to predictions of the “hygiene hypothesis”. Placebo tests using health outcomes that should not be affected by de-worming do not show similar patterns. Overall, childhood de-worming leads to improvements in morbidity and lifespan decades later.

## 1 Introduction

Over 400 million people globally live with hookworm (*1*). Seminal work finds human capital and short-term health benefits from mass de-worming (*2*, *3*). However, the value of large-scale de-worming continues to be debated, and long-term evidence on the consequences of de-worming is thin (*4*, *5*). This study contributes rigorous evidence on the adult lifespan and morbidity consequences of childhood de-worming, using the Rockefeller Sanity Commission’s (RSC) de-worming campaign in the US South as a natural experiment.

I also use de-worming to test the pathways linking early life infectious disease exposure and adult lifespan. While in-utero conditions have well-known consequences, less is known about long-term health impacts of childhood exposures (*6*). Reductions in early-life infectious disease has been implicated in 20th-century improvements in older-age mortality, and prior work links cohort infant mortality and respiratory disease prevalence to adult outcomes (*7* –*15*). Nonetheless, causal evidence and mechanistic understanding of the link between early life health and later life outcomes remain sparse. One challenge is that separating in-utero maternal exposure from childhood exposure is difficult. Additionally, infectious disease exposure may be endogenous to other socio-economic factors (*15*, *16*).^1^ This study’s approach addresses these concerns, leveraging rapid unanticipated reductions in hookworm prevalence that primarily affected children.

While prior work implicates lifelong improved inflammation and nutrition as mechanisms linking early life health and adult lifespan, mechanisms are rarely tested directly (*7*, *8*, *18*, *19*). Further, it is not theoretically clear that reduced infectious disease burdens are uniformly positive. The “hygiene hypothesis” suggests pathogen, particularly helminth, exposure may play a distinctive beneficial role in immune regulation (*20*, *21*). Helminth reductions have been linked to the rise of allergies and auto-immune conditions, but murine and human evidence is discrepant, theoretical mechanisms are contested, human evidence is mixed, and models and methods to critically evaluate the hygiene hypothesis have been limited (*21* –*23*).^2^ This paper finds long-term physiologic benefits of de-worming, with implications both for the mechanistic link between early life infectious disease, and for the role of hookworm in the hygiene hypothesis.

I study the RSC mass-deworming campaign, using an approach similar to Bleakley, 2007’s study of de-worming on schooling (*3*). Hookworm, primarily *Necator Americanus*, in the early 20th century United States was principally a disease among children that caused very little acute mortality, but potentially serious chronic morbidity and inflammation (*22*, *29*). In 1909, the RSC was established to eradicate hookworm in the American Southeast, with de-worming at scale by 1913 (*30*). De-worming was pharmaceutical using thymol, but the RSC developed systems, infrastructure, and education to enable continued identification, treatment, and prevention of hookworm after the campaign ((*30*)). Pre-campaign hookworm prevalence from the Rockefeller Archive is plotted in figure 1 (*31*). Nearly 40% of children had hookworm, with substantial geographic variation (*32*). Rapid, unanticipated reductions in hookworm due to the RSC campaign provides a natural experiment to explore the long-term health effects of hookworm.

**Figure 1:**
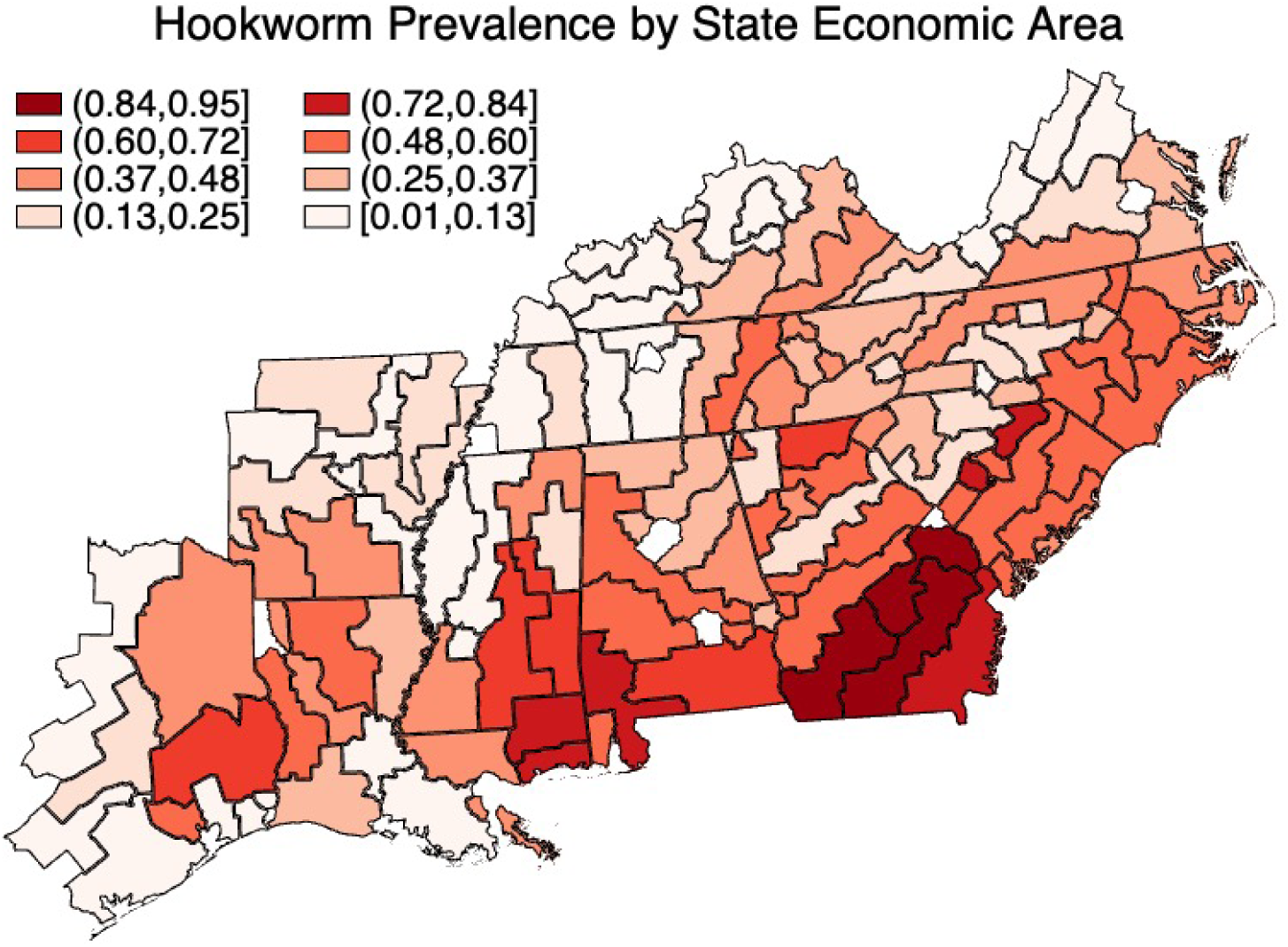
Distribution of pre-intervention hookworm by state economic area *Notes*: This figure shows estimates of childhood hookworm prevalence from systematic county surveys of children pre-intervention conducted by the Rockefeller Sanitary Commission. County data are aggregated and displayed at the State Economic Area (SEA). Prevalence data were digitized by Roodman, 2018 from the Rockefeller Foundation Archive (*31*).

My empirical approach compares individuals born in areas with varying baseline hookworm exposure, before and after the RSC campaign. I examine the impacts of de-worming before age 5 on lifespan in a sample of nearly 4 million older adult deaths drawn from Social Security Administration records. Importantly, I am able to examine variation across counties, but within states, and account for time-specific changes in each state. In a parallel set of analyses, I draw biomarker outcomes from nationally-representative samples, including immunologic processes such as allergy skin tests and erythrocyte sedimentation rate (an inflammation biomarker), other hookworm-related outcomes including BMI and hemoglobin, and a set of placebo health outcomes that are unrelated to hookworm exposure. This study provides rigorous evidence on the long-term impacts of hookworm on lifespan, as well as mechanistic insight into the ways hookworm may shape long-term health.

## 2 Results

### 2.1 Study populations

This study draws data from two populations. The primary lifespan analyses draw nearly 4 million death records from the Berkeley Unified Numident Mortality Database (BUNMD) (*33*, *34*). The BUNMD is derived from a 2013 transfer of Social Security Administration Numident mortality records to the National Archives and Records Administration. In primary analyses, I draw data from deaths between 1988-2005, the BUNMD “high coverage” years where over 95% of older adult deaths in the United States are included. I relax this restriction and evaluate earlier death years with lower mortality coverage as a robustness check. I also use census data to verify that selection into the sample is unrelated to the RSC campaign (supplement table A1). The sample is weighted to reflect the universe of deaths in the human mortality database (HMD). Critically, for each decedent these data include county of birth, as well as age and date of death. For empirical models, I aggregate birth counties into State Economic Areas (SEAs) with stable borders over time. I restrict to decedents born between 1900-1935 in areas of the American South where the RSC was active and where I have pre-campaign SEA-level hookworm prevalence data. This yields an analytic sample of 3,980,291 individuals, 47% of whom are male, and 27% are Black (table 1).

**Table 1:** Summary statistics by sample.

|  | **Unified Numident**<br>Decedents | **NHANES**<br>Survey Participants |
| --- | --- | --- |
| **Lifespan (years)** | 77.6<br>(7.2) | -<br>- |
| **Age (years)** | - | 57.18<br>(10.69) |
| **Proportion Male** | 0.47<br>(0.50) | 0.46<br>(0.50) |
| **Proportion Black** | 0.27<br>(0.45) | 0.15<br>(0.35) |
| **Effective Years** | 2.92<br>(1.34) | 2.34<br>(1.85) |
| **Proportion born in RSC State** | 1<br>- | 0.34<br>(0.47) |
| **Proportion in Major Urban Areas** | - | 0.46<br>(0.50) |
| **Proportion in Rural areas** | - | 0.39<br>(0.49) |
| **Proportion HS Grad+** | - | 0.47<br>(0.50) |
| **Proportion College Grad+** | - | 0.08<br>(0.28) |
| **Proportion low income** | - | 0.24<br>(0.43) |
| **N** | 3,980,291 | 18,377 |

Notes: This table presents summary statistics for individuals in both of the analytic samples used for these analyses. Mortality data are drawn from the Berkeley Unified Numident Mortality Dataset. Morbidity data are drawn from the NHANES I and NHANES II. Individuals represented were born between 1900 and 1935.

The distribution of birth years smoothly covers my birth cohorts of interest, the distribution of death years increases slightly over the years covered (Supplement figures A1,A3) (*33*).^3^ The average lifespan in this sample is 77.6, somewhat older than the expected lifespan for these birth cohorts, since my primary analysis of the BUNMD conditions on survival until the latter 20th century.^4^ It is important to consider that estimates are in a sample of older-age deaths. Selection into the sample on the basis of older ages of death may bias estimates toward 0 if additional individuals who died earlier than the period of observation due to hookworm were less healthy than those who survived into the observation window. I conduct additional analyses with census data and expanded samples to validate results.

My second sample draws from two waves of the National Health and Nutrition Examination Survey (NHANES), which were fielded in 1971-74, and 1976-80. Both surveys are nationally-representative probability samples of the United States, and include state of birth. I focus on those born from 1900-1935, for a sample of 18,377. Unlike the lifespan specification, I use state of birth to assign hookworm prevalence, and thus make comparisons across states. I restrict to individuals born in the contiguous United States, rather than just RSC-active areas. Morbidity outcomes are assessed at an average age of 57.2. 46% are male, 15% are Black, 39% live in rural areas, and 47% finished high school. Results should be interpreted as morbidity in mid-to-late life. I test several biomarkers from this population. I examine erythrocyte sedimentation rate (ESR), a biomarker for background inflammation; number of skin-tested allergies out of 8 skin pricks; body-mass-index, hemoglobin, and a series of placebo outcomes. Some outcomes were only measured in random sub-samples of the population.

### 2.2 Impacts of de-worming on lifespan

I evaluate the impacts of de-worming before age 5 on lifespan in the Unified Numident dataset using a primary specification defined in equation (1) that regresses lifespan on the interaction between baseline SEA-level hookworm prevalence and effective years of exposure, with fixed effects by sex for birth SEA, and state of birth-by-year of birth. SEA fixed effects absorb time-invariant SEA characteristics (including baseline hookworm prevalence and time-invariant differences such as economic characteristics and other disease environment), and state-by-birth year fixed effects capture potential state-specific trends. Thus, variation in pre-campaign exposure to hookworm across SEAs, within states, is used to estimate long-term impacts on lifespan.

I find one year of de-worming is linked to 0.042 additional life years (0.504 months) (Table 2). This effect is qualitatively large - the full implied impact of 5 years of childhood deworming exposure is 0.21 additional years of life at older ages, similar to the life expectancy gained by eliminating Alzheimers’ disease, half of cerebrovascular disease in older age, or removing lead in-utero (*35*, *36*).

**Table 2:** Change in lifespan per year of exposure to de-worming campaign.

|  | Lifespan (years) |  |  |
| --- | --- | --- | --- |
|  | All | Males | Females |
| **A. Primary specification** |  |  |  |
| **SEA Prevalence × Effective Exposure Years** | 0.042***<br>[0.012] | 0.034**<br>[0.015] | 0.046***<br>[0.016] |
| **Constant** | 77.765***<br>[0.012] | 76.100***<br>[0.016] | 79.191***<br>[0.015] |
| **N** | 3,980,291 | 1,851,939 | 2,128,352 |
| **Implied effect of full exposure (years)** | 0.211 | 0.171 | 0.228 |
| **B. Allow for SEA-specific mean reversion** |  |  |  |
| **SEA Prevalence × Effective Exposure Years** | 0.042***<br>[0.012] | 0.035**<br>[0.015] | 0.045***<br>[0.017] |

Notes: This table presents estimates of the effect of exposure to the de-worming campaign on the age of death, first pooled then stratified by sex. Lifespan data are drawn from decedents in the CenSoc Unified Numident file (BUNMD), and include deaths in high-coverage years from 1988-2005. The sample includes decedents born between 1900 and 1935 in the 11 states the RSC was active, in the SEAs where pre-intervention surveys were conducted. Hookworm prevalence is assigned at the SEA of birth. Estimates are conducted using fixed effects by sex that reflect birth SEA, and birth state-by-birth year. Standard errors presented in brackets, clustered at the SEA level. The coefficients presented are the interaction between baseline SEA hookworm prevalence and effective years of exposure, and can be interpreted as the implied effect of one full year of exposure to de-worming. Each individual received up to five years of exposure. Panel B shows the same model estimated allowing for SEA-specific mean reversion based off of SEA lifespan in the year prior to the campaign's initiation.

The point estimates for females are larger than for males, though the difference is not statistically significant. This is potentially consistent with the fact that women have conventionally benefited more than men from improvements in the infectious disease environment (*37*).

Two event-study style analyses support the argument that changes in lifespan are driven by de-worming due to the RSC-campaign. In the top panel of figure 2, an event-study with a full set of fixed-effects from the primary specification examines the change in the estimated relationship between baseline hookworm prevalence and lifespan within each birth cohort, relative to the last birth years before the intervention began. In the 12 preceding years, there is no evidence that the outcomes for areas with more hookworm prevalence were evolving differently from those with less, supporting the validity of the parallel-trends assumption. After the RSC campaign, the coefficient changes positively, suggesting increases in lifespan related to de-worming that occur shortly after campaign initiation. On the bottom panel, a similar analysis is conducted, but the SEA fixed effects are excluded, so the baseline hookworm prevalence-lifespan relationship can be estimated directly for each birth cohort. A similar pattern is visible - there are no systematic trends related to hookworm prevalence in the pre-period. In the post period, the coefficient goes from negative to essentially 0, suggesting that the relationship between baseline hookworm prevalence and lifespan did not just become more positive, but that large-scale treatment of hookworm rendered baseline hookworm prevalence and lifespan uncorrelated after it occurred.

**Figure 2:**
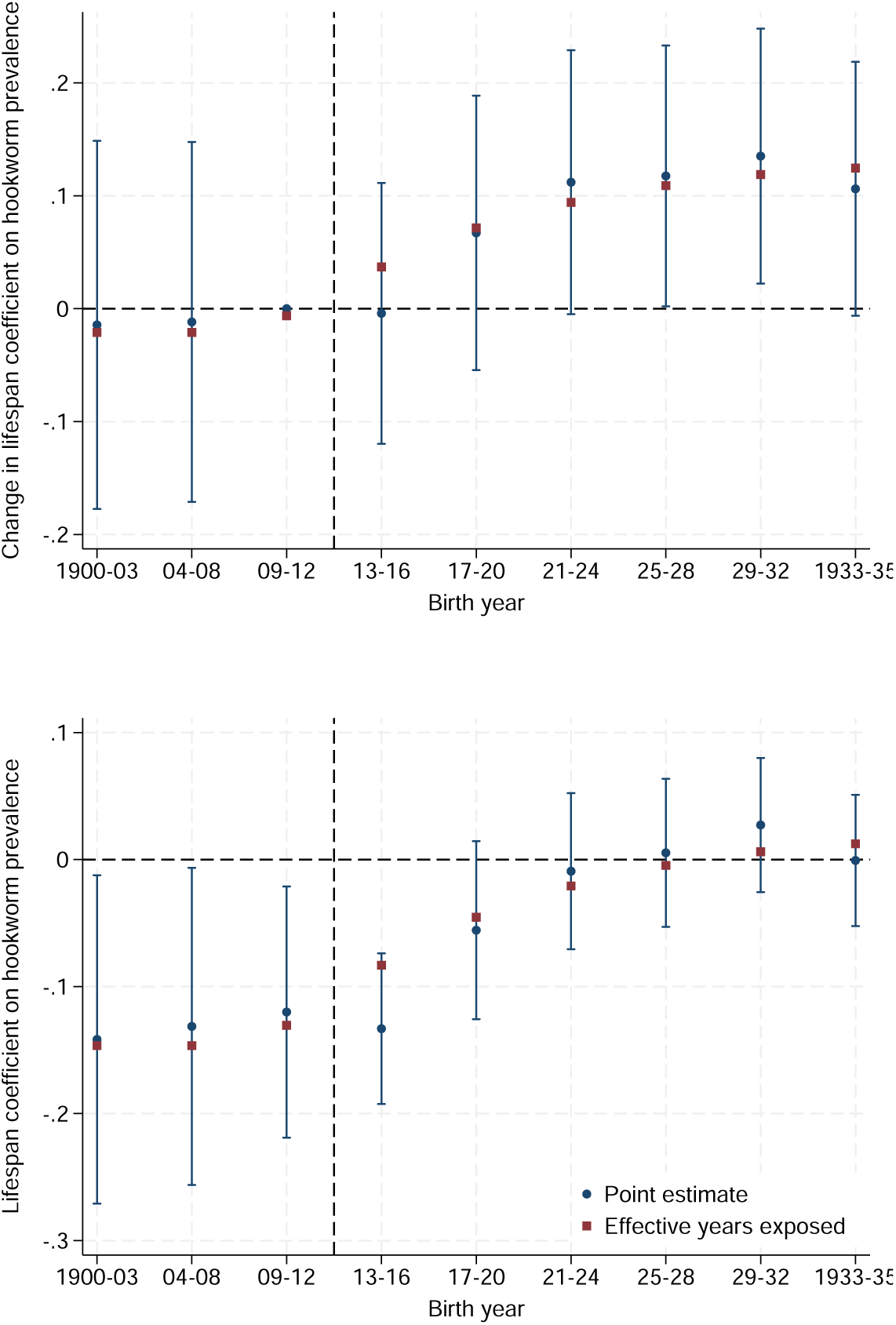
Relationship between baseline hookworm prevalence and lifespan across cohorts *Notes*: This figures show estimates of the relationship between *baseline* state economic area (SEA) hookworm prevalence and lifespan for people born in a given SEA, estimated for different birth cohorts. The top panel plots event study estimates, including fixed effects by sex that reflect birth SEA, and birth state-by-birth year. Plotted coefficients are the interaction term between birth year and hookworm prevalence, reflecting the hookworm prevalence coefficient in each birth cohort, relative to the years immediately pre-campaign. The bottom panel relaxes this model, dropping the birth SEA fixed effects. Thus, the relationship between baseline hookworm prevalence and lifespan can be directly estimated and plotted for each birth cohort. Blue markers plot coefficient estimates and 95% confidence intervals within each cohort over time, visualizing the trajectory of the changing relationship between baseline hookworm prevalence and lifespan around the time of RSC campaign initiation. Red squares show the normalized average effective years of exposure in each set of year bins, to map plotted results onto the functional form applied in the primary estimate in table 2. In order to visualize the functional form of effective years of exposure assigned alongside the point estimates, I regress the effective years on the point estimate, and plot the normalized values. Detailed effective years for each birth cohort are plotted in supplement figure A5.

Results are robust to accounting for potential mean reversion.^5^ Results are also robust to a variety of alternative specifications and functional forms including linear years of exposure before age 5, a binary pre/post specification, and a semi-parametric specification using tertiles of baseline hookworm prevalence, as well as including controls for birth year interacted with rurality, climate, and soil suitability for cultivation from the FAO-GAEZ (supplement tables A2,A3).^6^

A potential concern is that these data do not record lifespans for deaths that occur earlier in the 20th century. If hookworm led additional individuals to die earlier than the period of observation, and those individuals were less healthy than those who would have survived anyways, the effect I find is an underestimate of what one would find if all deaths were observed. Nonetheless, I conduct parallel analyses of survival until 1960 using state data from the census, focusing on the same RSC-campaign states. I find de-worming significantly increases the probability of survival until 1960 (supplement table A4). I also estimate the impacts of de-worming using a broader sample of the Unified Numident data that expands coverage primarily to younger ages of death (5.3 million obs.), including years before 1988 with lower mortality coverage, and extending coverage to 2007. Findings are very similar, but point estimates are larger, suggesting that the primary estimates from this paper may be underestimating total potential impacts (supplement table A5).^7^

### 2.3 Impact of childhood de-worming on morbidity

Using data from the NHANES I and NHANES II, I estimate a parallel model to the specification for lifespan, using birth-state hookworm prevalence pre-campaign in the contiguous United States. I focus on domains in which childhood exposure to hookworm may persist: health outcomes related to immunologic changes, and outcomes related to nutritional status and anemia.

I find evidence for long-term immunologic improvements related to de-worming (table 3). Estimating the effect of 1 year of de-worming on the number of skin-tested allergies finds reductions of 0.14. The implied reduction in the number of allergies is substantial: five years of childhood de-worming exposure is estimated to reduce the average number of allergies by nearly 0.75, approximately 1/2 of a standard deviation. I also find each year of de-worming reduces erythrocyte sedimentation rate, a biomarker for inflammation, by 1.5mm/hr later in life, which is significant at a 10% size of test. This implies a reduction by 7.5mm/hr with full campaign exposure ( 3/4 of a standard deviation). This is slightly larger than the gap between people born pre-intervention in the South versus the rest of the country.

**Table 3:**
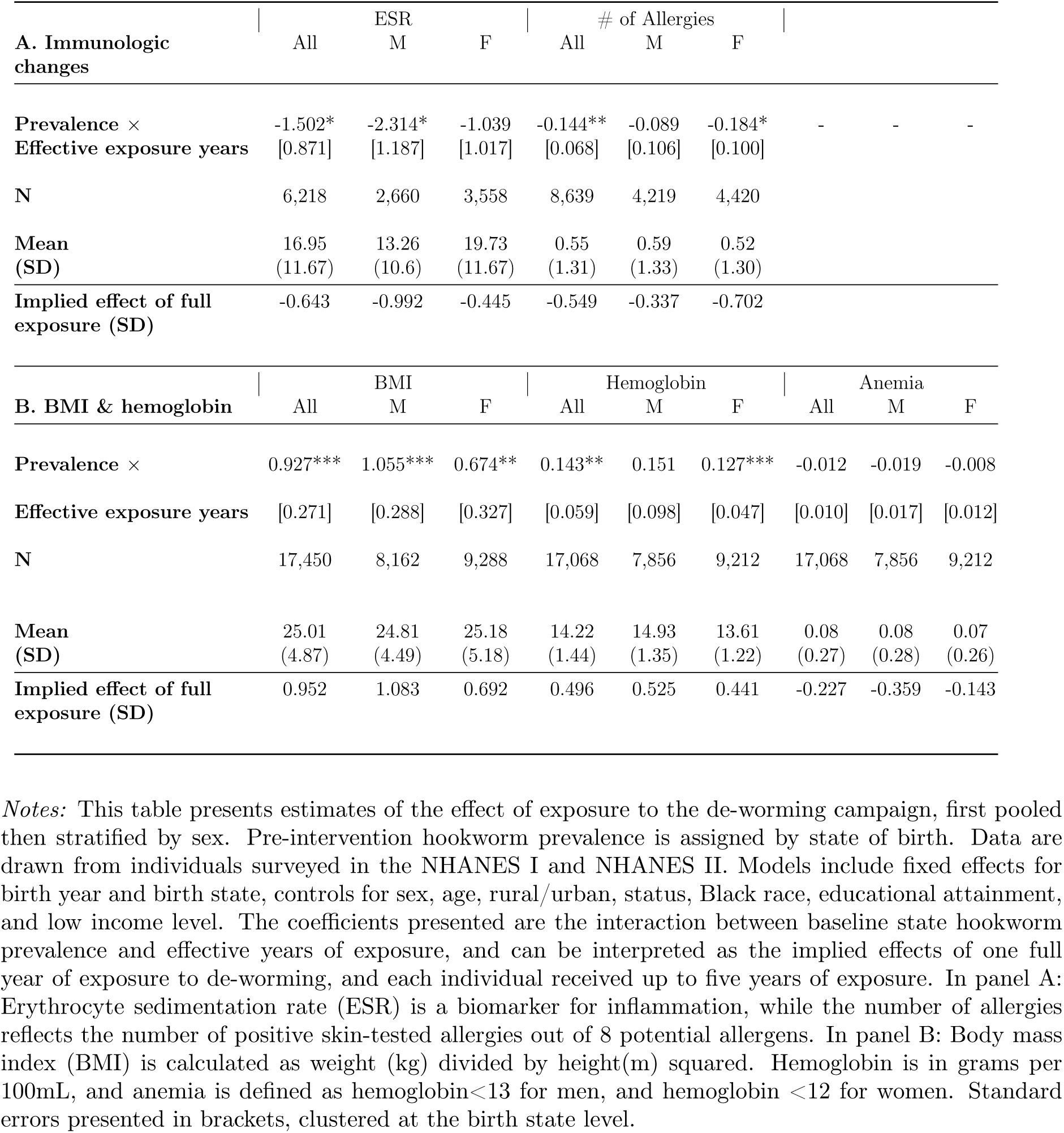
Estimated change in adult morbidity per year exposure to de-worming (before age 5).

Beyond its immunologic impacts, hookworm also may cause decreases in BMI and hemoglobin, which may be persistent. I find that exposure to de-worming is linked to increased BMI and hemoglobin in adulthood, with full exposure associated with nearly a 1 standard deviation increase in BMI, and a nearly 1/2 standard deviation increase in hemoglobin. This appears to be true for both males and females. The increases in BMI are apparent across the BMI distribution, but de-worming appears to especially reduce incidence of low BMI and increase sub-obese overweight (Supplement table A6). The benefits for hemoglobin appear to be sub-clinical improvements in hemoglobin status. While the point estimates for anemia are negative, they are small and not statistically significant.

In the NHANES data, unlike the lifespan data, I can control for individual-specific socioeconomic and geographic characteristics at the time of measurement ^8^. I include these in the estimates shown, but estimates are minimally-affected when a simpler specification without controls is used.

### 2.4 Placebo health tests

I examine health outcomes that are unlikely to be significantly affected by hookworm, in order to test whether other health changes occurring at the same time as the RSC campaign and spuriously correlated with de-worming — but not caused by it — might compromise the identification of de-worming’s potential long-term impacts (table 4).

**Table 4:**
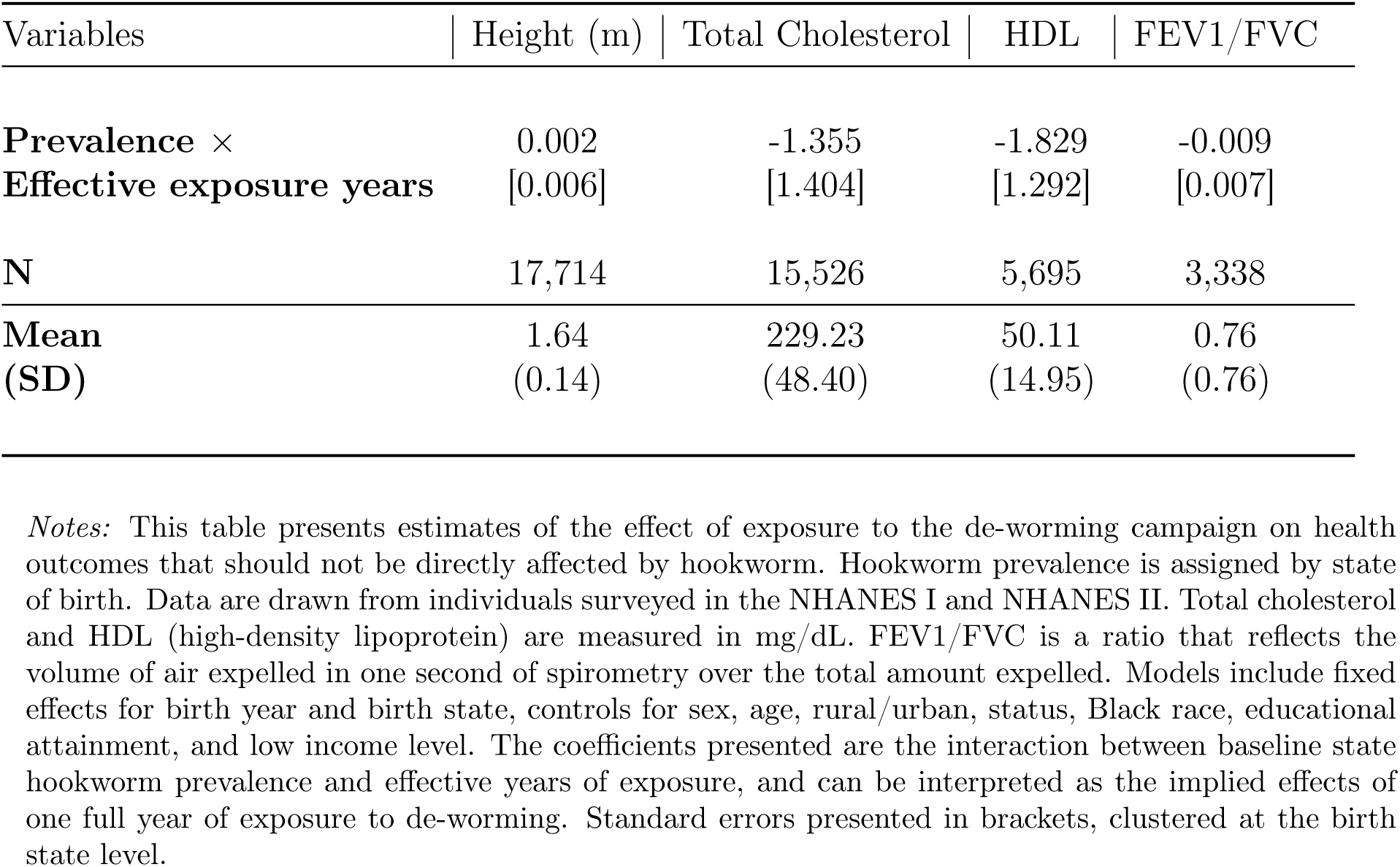
Placebo tests.

| Variables | Height (m) | Total Cholesterol | HDL | FEV1/FVC |
| --- | --- | --- | --- | --- |
| **Prevalence ×**<br>**Effective exposure years** | 0.002<br>[0.006] | -1.355<br>[1.404] | -1.829<br>[1.292] | -0.009<br>[0.007] |
| **N** | 17,714 | 15,526 | 5,695 | 3,338 |
| **Mean**<br>**(SD)** | 1.64<br>(0.14) | 229.23<br>(48.40) | 50.11<br>(14.95) | 0.76<br>(0.76) |

*Notes:* This table presents estimates of the effect of exposure to the de-worming campaign on health outcomes that should not be directly affected by hookworm. Hookworm prevalence is assigned by state of birth. Data are drawn from individuals surveyed in the NHANES I and NHANES II. Total cholesterol and HDL (high-density lipoprotein) are measured in mg/dL. FEV1/FVC is a ratio that reflects the volume of air expelled in one second of spirometry over the total amount expelled. Models include fixed effects for birth year and birth state, controls for sex, age, rural/urban, status, Black race, educational attainment, and low income level. The coefficients presented are the interaction between baseline state hookworm prevalence and effective years of exposure, and can be interpreted as the implied effects of one full year of exposure to de-worming. Standard errors presented in brackets, clustered at the birth state level.

I first examine height in the NHANES. Height is not a perfect placebo outcome, as extreme calorie deprivation can reduce heights. However, most hookworm infections were not extreme, and height also reflects in-utero health, early life conditions, and characteristics of parents that may be changing in the background (*15*, *38*, *39*). While point estimates are small and positive, I find no significant effect of exposure to the de-worming campaign and height. This is particularly re-assuring insofar as it indicates little in the way of in-utero changes or other early-life changes.

I also examine cholesterol measures, which reflect not only health endowments but also health-related behaviors such as diet, exercise, and health care access. I find no impacts of de-worming on cholesterol measures.

Finally, I find no effects of de-worming on FEV1/FVC, a common pulmonary function test that is sensitive for asthma.

## 3 Discussion

I examine the impact of de-worming before the age of 5 on adult lifespan and morbidity later in life. In doing so, I provide evidence on the long-term impacts of de-worming, as well as novel tests of 1) the linkage between early-life inflammatory exposure with later life morbidity and mortality, and 2) the role of hookworm in the “hygiene hypothesis” in the 20th century. I find substantial gains in lifespan at older age, as well as long-term reductions in a biomarker of inflammation and the number of skin-tested allergies, as well as increases in BMI and hemoglobin. Similar changes are not observed in health outcomes that should be unaffected by de-worming.

The estimated impact of de-worming on lifespan at older age is large, corresponding roughly to the older-age lifespan lost due to Alzheimer’s Disease. These findings are consistent with, and provide new rigorous evidence for, influential theories that infectious disease burdens early in life shape mortality across the life course (*7*, *18*). Men and women benefit similarly from de-worming, though point estimates are larger for women. Sex differences in epidemiological and laboratory studies of immunology are becoming increasingly appreciated, in part improvements in women’s lifespan relative to men over the 20th century have been attributed to this (*37*, *40*, *41*). The morbidity findings provide mechanistic support for many of the theorized links between early life infectious disease and adult lifespan. Changes in ESR are consistent with long-term changes in chronic, low-grade, inflammation that may raise cardiovascular disease and cancer risk (*42*, *43*). Further, changes in early-life health such as nutritional status and hemoglobin may be persistent, leading to long-term benefit (*8*, *44*). In this sample of older adults with relatively low BMI in the 1970s, I interpret increases in BMI as likely positive protections against frailty, especially as changes are concentrated among reducing low weight and sub-obese BMI (*45*, *46*).

While the improvements in mortality can be attributed to meaningful observed improvements in underlying health changes from early childhood, it is possible that de-worming may lead to economic benefits that may also play a role (*2*, *47*). While I do not have educational attainment and mortality data in the same individuals, it is unlikely that changes in education or income fully explain the observed impacts. In a study of the economic ramifications of under-18 exposure to de-worming, Bleakley finds no long-term impact on educational attainment, but small increases in income. In my analyses of morbidity, I directly control for a variety of potentially relevant socio-economic pathways including education, poverty status, and rurality. These controls minimally impact observed results.

The joint reductions in background inflammation as well as allergies further an argument that helminths may in fact raise allergies through chronic inflammatory processes (*23*). Notably, these findings contrast sharply with a potential role of hookworm in the “hygiene hypothesis” (*20* –*23*, *25*, *48*). This study provides rigorous evidence of long-term impacts between hookworm and allergies, in a space where mouse models and short-term human trials are conflicting, and active debate ensues. However, the interpretation of these findings is limited by the context of the RSC de-worming campaign in the early 20th century. The role of hookworm in auto-immune processes may vary in contexts with differing levels of background pathogens (*49*).

This study also has substantial implications for mass de-worming programs around the globe. The efficacy and cost-effectiveness of mass de-worming has been debated in recent years, with mixed results on the impacts of hookworm on short and medium-term outcomes, and theoretical arguments made that the potential role of hookworm in auto-immune conditions may undermine the value of de-worming (*4*, *21*, *50*). Prior work on the RSC campaign has found long-term improvements in income and contemporaneous improvements in school attendance (*3*), and contemporaneous work on male deaths finds health impacts of deworming and education may be complementary for adult lifespan (*51*).^9^ This study provides evidence for life-long benefits to de-worming, with persistent impacts on health and age at death. These benefits make a strong case for expanded de-worming programs in endemic areas around the globe.

This study has several limitations. One limitation is that the primary source of mortality data primarily draws from the later 20th century, potentially missing earlier-life impacts of hookworm. Hookworm should minimally impact childhood mortality, but may have impacted mid-life mortality, which would not be measured in this paper’s primary analysis. Analyses at the state level using census data on survival rates until 1960, which should reflect mid-life mortality, as well as extensions of the Unified Numident sample into earlier years of death, find larger impacts of hookworm exposure relative to the primary sample. Primary estimates are thus possibly underestimates of true lifespan effects.

Another potential limitation is unobserved changes in the health status across cohorts before and after de-worming begins, in a way that is related to hookworm prevalence. I address this in several ways. 1) Using a model that includes state-by-birth year fixed effects in the primary mortality specification, I am able to account for potential state-specific trends and changes over time, and localize the variation used to across-SEAs within states. I also extend the model to include controls for time trends that may be related to SEA characteristics such as rurality, climate, and soil suitability for agriculture. 2) Event-study analyses suggest that in the years before the campaign, and the years after full effects are realized, mortality dynamics are not evolving in a way related to baseline hookworm prevalence. 3) I provide evidence from placebo health outcomes that there were not major changes in health status unrelated to hookworm-specific morbidity.

I conclude that de-worming in early childhood has large life-long benefits for morbidity and mortality. Mechanistic evidence finds support for long-term immunologic improvements due to de-worming, and other health benefits. In this context, hookworm reduces allergies in a way that is not consistent with a role in the “hygiene hypothesis”. Potential long-term benefits should be considered when evaluating mass de-worming programs.

## 4 Methods

The intuition for these analyses is analogous to a difference-in-differences approach, similar to Bleakley, 2007. While conventional difference-in-differences uses a treatment variable that takes the value of 1 for groups eventually treated, and 0 untreated, I use continuous hookworm prevalence from prior to the intervention that takes values between 0 and 1. Individuals from areas with higher hookworm prevalence should disproportionately benefit from the de-worming campaign. Comparing birth cohorts before and after the de-worming campaign from areas of varying hookworm prevalence enables a causally-interpretable estimate of the long-term consequences of childhood de-worming.

### 4.1 Data sources for hookworm prevalence

Prior to the initiation of the de-worming campaign, the RSC sampled at least 200, but often many more, children in each county. They examined stool microscopically for hookworm ova. Using data digitized by Roodman from the Rockefeller foundation archives, I aggregate counties into State Economic Areas (SEAs) with stable borders over time (*29*, *31*). As shown in figure 1, prevalence varied within and across states before the campaign. Hookworm suitability was primarily a function of land characteristics and temperature: sandy soils were preferable for hookworm reproduction. Note that many of the largest urban centers were excluded from the pre-intervention prevalence data and activity. People born in these areas are also excluded from analysis. Prior analyses of pre-intervention hookworm prevalence found more rural and less Black counties had higher prevalence, but the primary variation was predicted by soil type and number of frost-free days (*30*). I use this pre-intervention hookworm prevalence data in my primary mortality specification.

In other analyses, I take a complementary approach comparing hookworm prevalence across states. To expand my sample to the contiguous United States, I reference complementary hookworm data from army recruits (*52*). Analyses of the 11 overlapping states suggests that the supplementary dataset well-approximates systematic county-level data collection from the RSC (supplement figure A4). The primary difference is that army prevalence is lower, on average, than the county data for a given prevalence level. This is likely due to differences in the age of the samples.^10^

### 4.2 Defining exposure to the RSC campaign

In order to account for 1) partial treatment of children born right before the initiation of the RSC campaign, and 2) the fact that it took several years to achieve high penetrance of deworming, I construct a continuous measure of “effective years” of exposure to the campaign before age 5. To model the increasing efficacy of the campaign over time, I use aggregated data from a subset of counties where repeated follow-ups occurred. In these samples, in 1910 59.7% of children had hookworm, falling to 39.7 in 1915, to 21.7 by 1920. This suggests yearly reduction in hookworm prevalence by a factor of 0.9. The functional form is as follows: 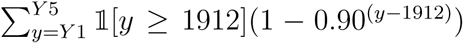 where *Y* 1*, Y* 5 are the years of birth through age 5 respectively. Thus, each individual gets the sum of “effective years” of exposure to the campaign before they reach the age of 5. The functional form is plotted in supplement figure A5. Analogous to more simpler approaches that use “years of exposure”, in the pre-period individuals have 0 effective years, and by 1920 the functional form is essentially flat. Results are robust to using simpler functional forms (A2).

### 4.3 Primary analyses of mortality

The empirical strategy rests on two core ideas. First, it leverages the varying hookworm prevalence across different areas at baseline. Higher prevalence areas stood much more to benefit from the RSC treatment than lower-prevalence. Second, the study design treats the initiation of the RSC campaign as an exogenous natural shock - given poor baseline understanding of hookworm and the amount of capital investment needed to intervene, the timing and initiation of the RSC campaign is considered exogenous to later health outcomes.

The core specification for the lifespan outcome is as follows:

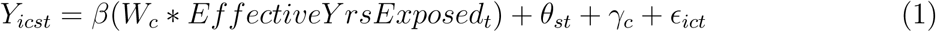

Where *Y_icst_* is lifespan for an individual *i* born in state economic area *c* and state *s*, in year *t*. *W_c_* refers to the hookworm prevalence at baseline in SEA indexed by *c*, and *EffectiveY earsExposed* is based on birth year and is the effective number of years exposed to the RSC campaign. *θ_st_* represents the state of birth-by-year fixed effects, which capture state-specific shocks and time-trends. *X_i_* represents covariates of interest - while a general label is used, in lifespan analyses the only covariate used is sex. *γ_c_* are fixed effects for SEA *c*. These terms are allowed to vary by sex as potentially important sex-specific trends may have been different for males and females during this time (*53*, *54*). In regressions separated by sex, these effects collapse to just one term for birth SEA, state, or year.

Note, *γ_c_* absorbs the baseline worm prevalence, along with observed and unobserved SEA-level characteristics that may be relevant for lifespan and are not changing over time. While there are some concerns for this study about limited coverage of deaths at younger ages and selection into the sample, the birth state by birth year fixed-effect capture cohort-level endogeneity in the probability a given individual is reflected in the data, to the extent it is fixed across a given cohort. *θ_st_* also accounts for observed and unobserved state and cohort effects in lifespan. The analytical sample is restricted to RSC-surveyed SEAs.

Taken together, the coefficient of interest *β* can be interpreted as the effect of an additional year of exposure to de-worming on age at death, estimated by comparing across SEAs, but within states, that have differing baseline prevalence of hookworm. Standard errors are clustered at the SEA level.

### 4.4 State-level survival estimates

In order to address concerns that the primary mortality data are at older ages and may miss earlier-life mortality, I conduct a second series of analyses using survival rates I construct from census data. In brief, I construct survival rates until 1960 from the census for birth cohorts from 1900-25, where the numerator is population in 1960 and the denominator the population alive at ages 5-14. I use population observations after age 5 for the denominator to avoid infant mortality. Using these state level survival rates, I estimate a similar approach with state-level hookworm prevalence, and a state fixed effect instead of SEA.

### 4.5 Morbidity analyses

I use a parallel strategy to the primary analyses of lifespan, in order to evaluate the impacts of de-worming on morbidity. I use a series of biomarkers from the NHANES. Since the NHANES does not have county or SEA of birth, I construct treatments at the state of birth, using state level hookworm prevalence from Kofoid and Tucker, 1921.

The core specification for each morbidity outcome is as follows:

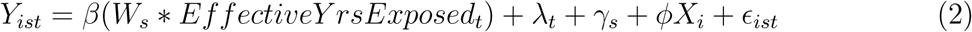

Where *Y_ist_* is the outcome of interest for an individual *i* born in state *s* in year *t*. *W_s_* refers to the hookworm prevalence at baseline in states indexed by *s*, and *EffectiveY earsExposed* is based on birth year and is the effective number of years exposed to the RSC campaign. *λ_t_, γ_s_* are birth state and birth-year fixed effects, which capture time-invariant state characteristics, and cohort-specific effects. These terms are allowed to vary by sex. One strength of the NHANES is richer socio-economic data. I include *X_i_* as a vector of covariates of interest, including sex, NHANES wave, age, rural/urban status, Black race, educational attainment, and low income level. Note that *γ_s_* will absorb baseline state hookworm prevalence, along with observed and unobserved state-level characteristics. *λ_t_* will absorb birth year-specific effects. *β* retains a similar interpretation to the primary specification, as the effect of an additional year of exposure to de-worming on lifespan, estimated by comparing across states that have differing baseline prevalence of hookworm, before and after the de-worming campaign. Standard errors are clustered at the state level.

## Data Availability

All data are public and available online:

https://wwwn.cdc.gov/nchs/nhanes/nhanes1/default.aspx

https://censoc.berkeley.edu/data/

## Acknowledgments

Many thanks to David Cutler, Ellen Meara, Lawrence Katz, Duncan Thomas, Claudia Goldin and Kirti Nath for helpful comments and discussion in the development of this draft. This work was supported by the National Institute of Aging (T32AG51108) and the National Institute of General Medical Sciences (T32GM144273).

---

**Figure A1:**
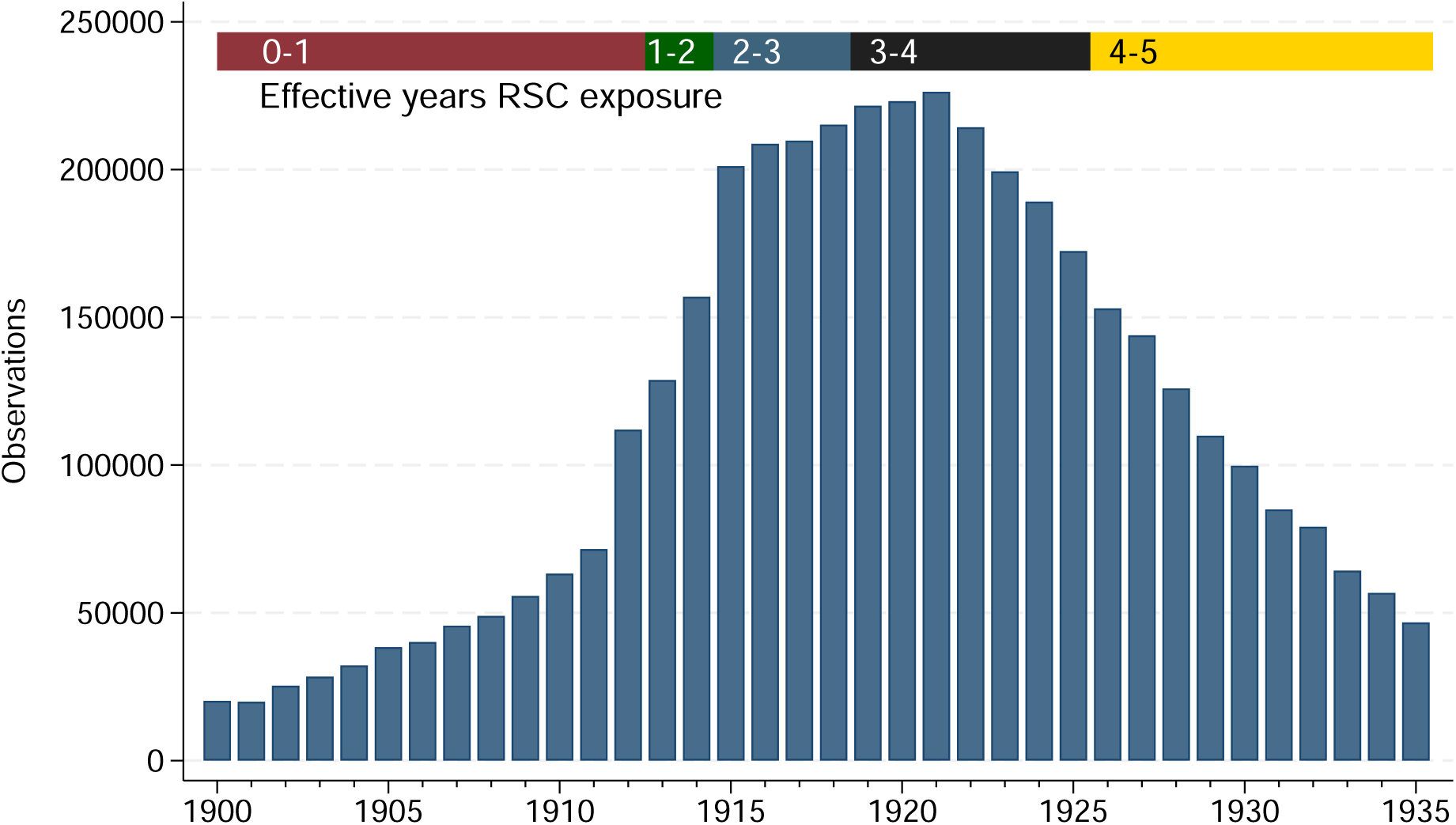
Distribution of birth years in the analytic sample *Notes*: This figure plots the count of individuals represented in the analytical sample by birth year. The top bar provides bands of the effective years of RSC campaign exposure that each birth cohort is assigned.

**Figure A2:**
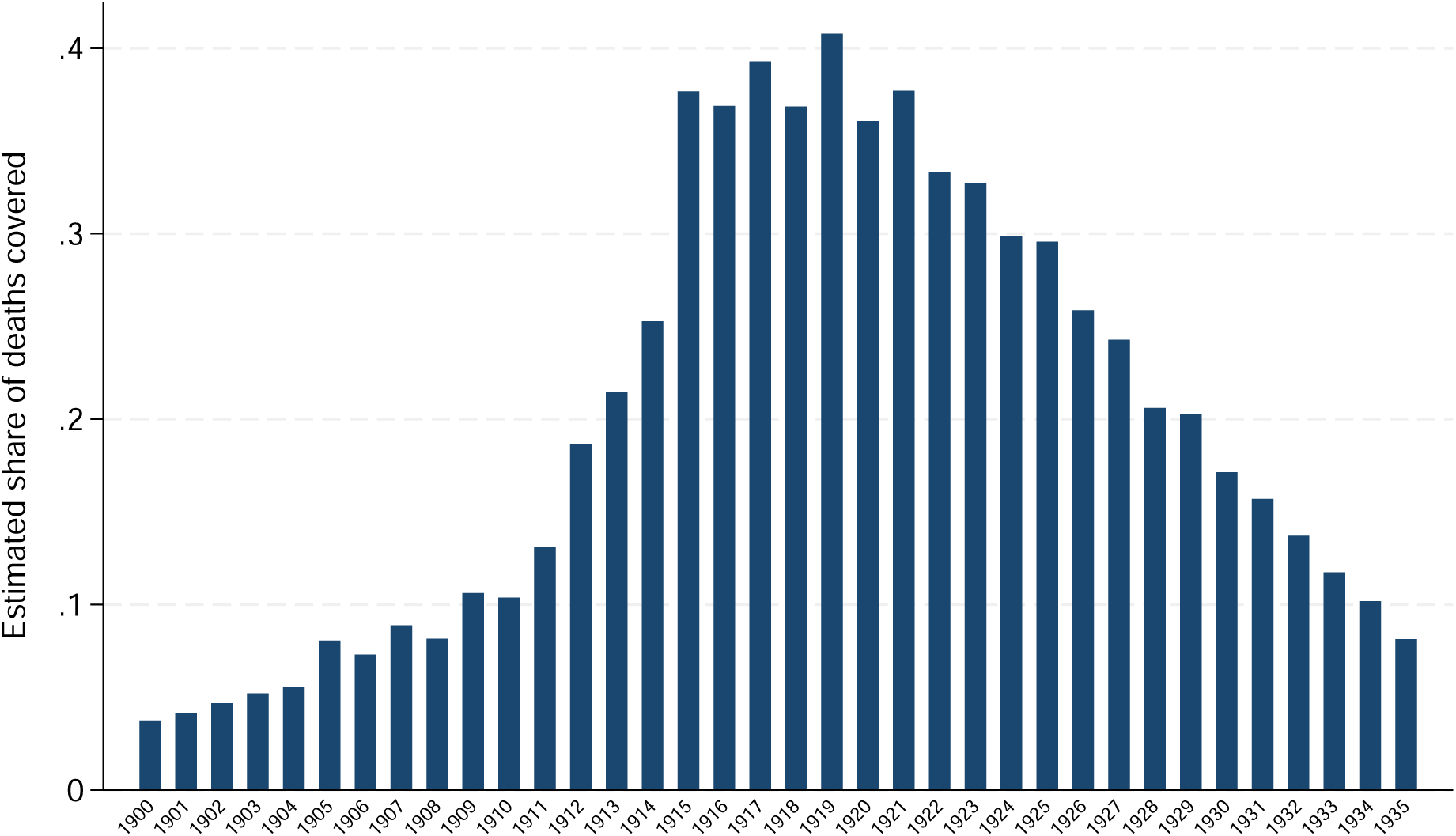
Estimated share of mortality covered of each birth year in the analytic sample *Notes*: This figure plots estimates of the share of deaths represented of each birth cohort in the analytic sample. Birth cohorts from 1900-1935 are included, and for each a proportion is produced where the numerator reflects the number of people born in a given birth-year-by-SEA that are in the high-coverage BUNMD, and the denominator reflects the number of people in an SEA from a given birth year that appear in the first census after age 5 a birth cohort is observed in (to avoid infant mortality). Each birth-year-by-SEA thus has one observation, weighted by population.

**Figure A3:**
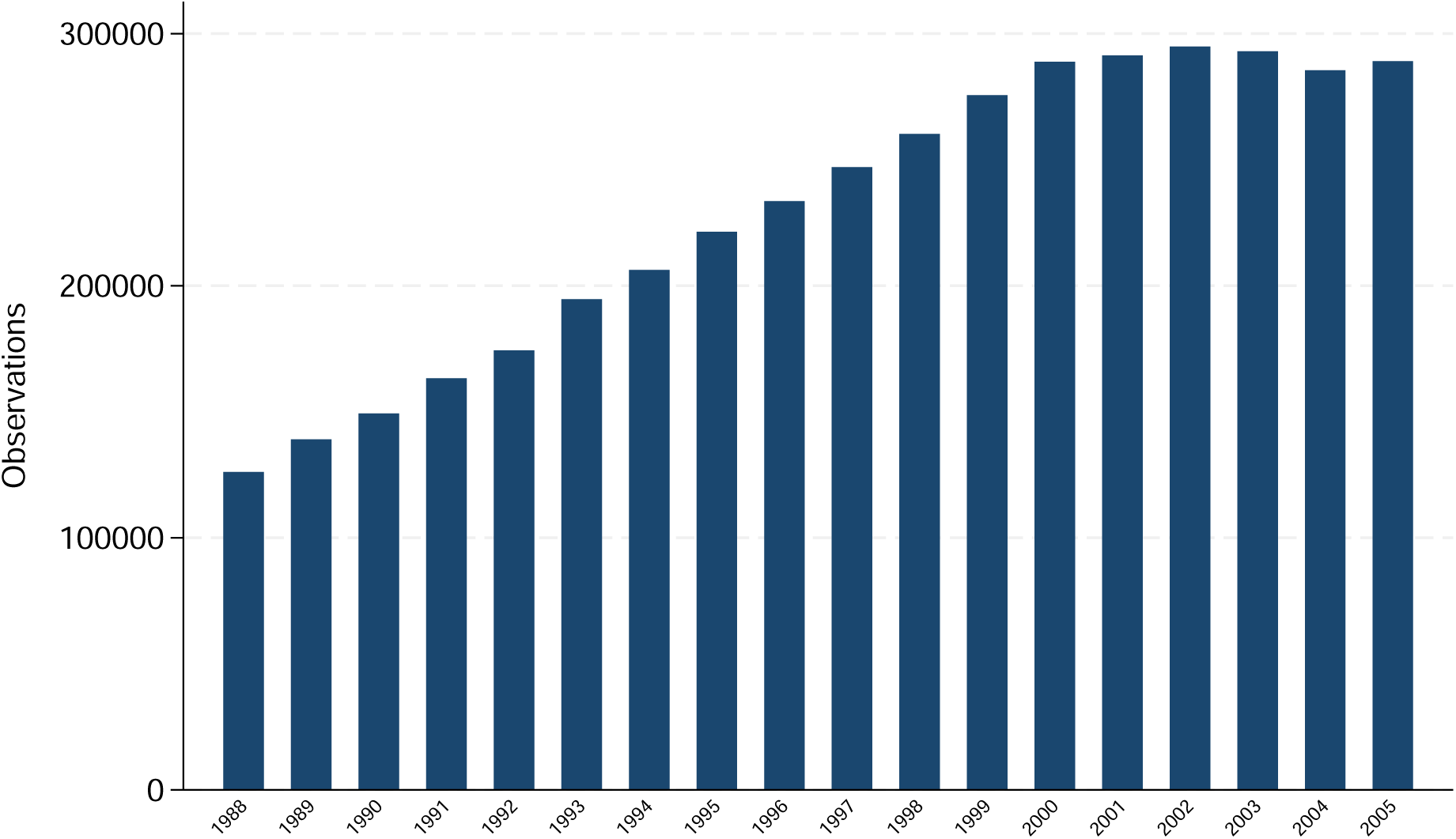
Distribution of death years in the analytic sample *Notes*: This figure plots the count of individuals represented in the analytical sample by death year.

**Figure A4:**
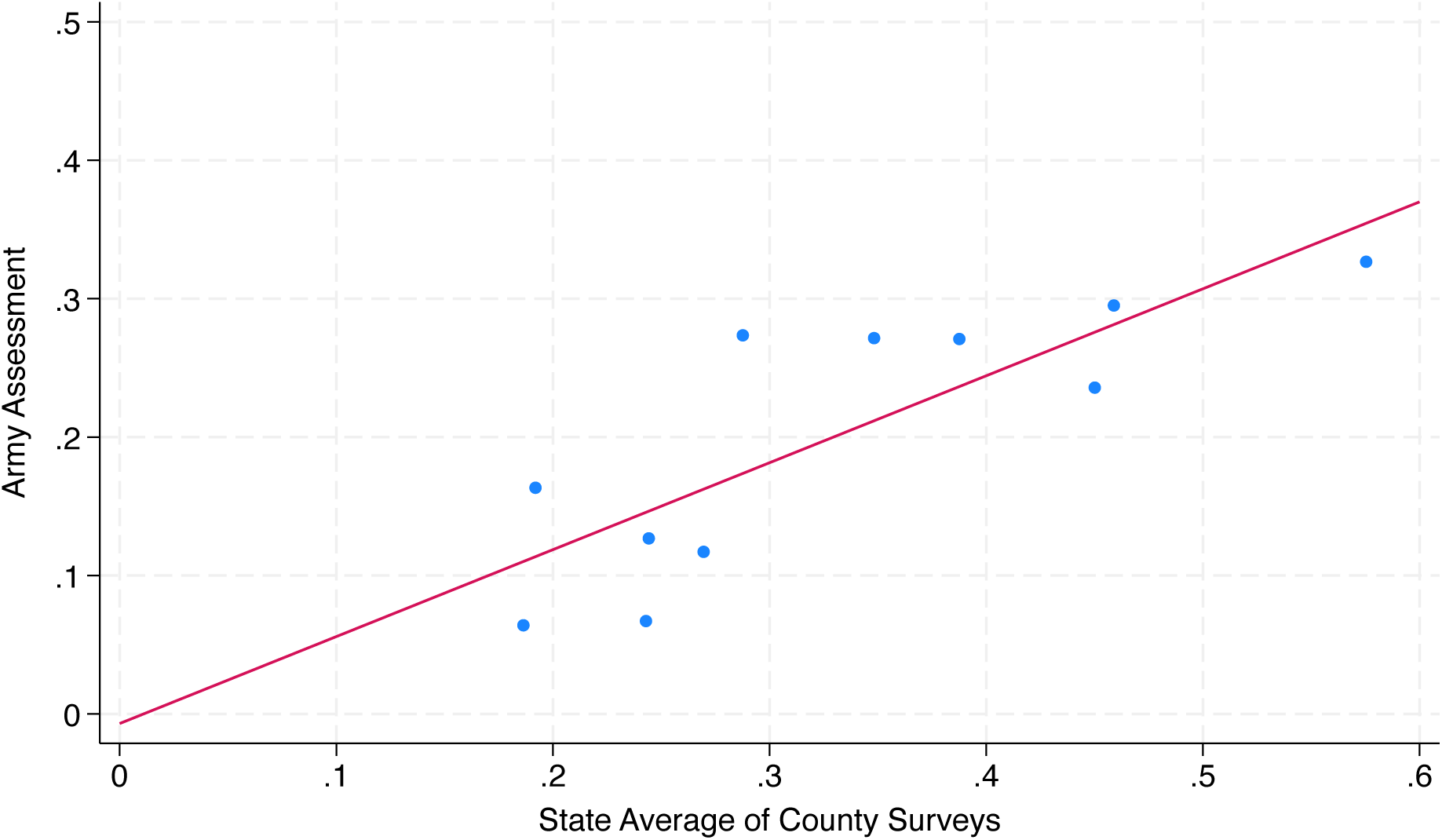
Relationship between army hookworm prevalence and county-level RSC assessed prevalence in overlapping states *Notes*: This figure shows the relationship between average hookworm prevalence from systematic RSC surveys of children in each county, and average hookworm prevalence from surveys of army recruits. The line of best fit is included.

**Figure A5:**
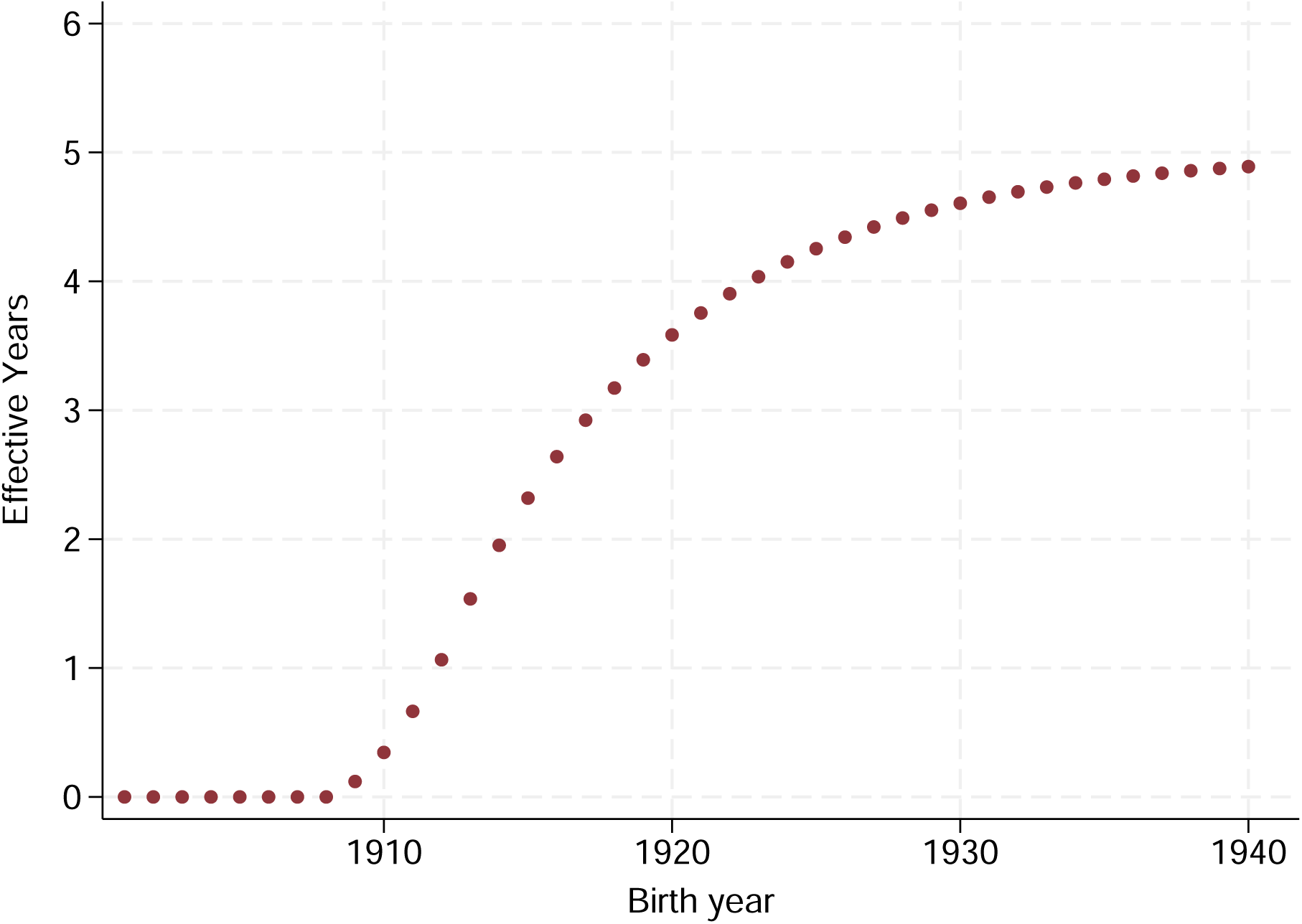
Effective years of exposure assigned to each birth cohort *Notes*: This figure shows the number of effective years of exposure assigned to each birth cohort over time.

**Table A1:** Evaluating whether representation in the BUNMD is related to the treatment of interest.

| **Fraction of individuals in Unified Numident** |  |
| --- | --- |
|  | **All** |
| **A. Primary specification** |  |
| **SEA Prevalence × Effective Exposure Years** | -0.001<br>[0.002] |
| **Constant** | 0.273<br>[0.002] |
| **N** | 4,212 |

*Notes:* This table presents an analagous estimate to the primary specification evaluating the impact of hookworm exposure, with an outcome that reflects the proportion of individuals observed in each census in an SEA that appear in the BUNMD. Birth cohorts from 1900-1935 are included, and for each a proportion is produced where the numerator reflects the number of people born in a given birth-year-by-SEA that are in the high-coverage BUNMD, and the denominator reflects the number of people in an SEA from a given birth year that appear in the first census after age 5 a birth cohort is observed in (to avoid infant mortality). Each birth-year-by-SEA thus has one observation, weighted by population. While the 1988-2005 years of the BUNMD reflect >95% of deaths among older adults in those years, this formally tests whether selection into the BUNMD sample is related to the RSC treatment. The specification reflects the primary specification using SEA fixed-effects, and state-by-birth year fixed-effects. Baseline hookworm prevalence is assigned at the SEA. Standard errors presented in brackets, clustered at the SEA level.

**Table A2:** Results from alternative specifications to estimate the impact of de-worming on lifespan.

|  | Longevity (years) |  |  |
| --- | --- | --- | --- |
|  | All | Males | Females |
| **A. Semi-parametric specification** |  |  |  |
| SEA tertile 1 × Effective Exposure Years (omitted) | - | - | - |
| SEA tertile 2 × Effective Exposure Years | 0.016**<br>[0.006] | 0.014*<br>[0.008] | 0.017**<br>[0.008] |
| SEA tertile 3 × Effective Exposure Years | 0.019***<br>[0.007] | 0.017**<br>[0.008] | 0.020**<br>[0.009] |
| Constant | 77.772***<br>[0.011] | 76.104***<br>[0.014] | 79.199***<br>[0.013] |
| N | 3,980,291 | 1,851,939 | 2,128,352 |
| **B. Pre-post specification** |  |  |  |
| Baseline SEA prevalence × [1] $>2$ effective exposure years | 0.112**<br>[0.048] | 0.049<br>[0.049] | 0.142**<br>[0.061] |
| Constant | 77.777***<br>[0.013] | 76.122***<br>[0.014] | 79.197***<br>[0.015] |
| N | 3,980,291 | 1,851,939 | 2,128,352 |
| **C. Linear years of exposure before age 5** |  |  |  |
| Baseline SEA prevalence × years since 1912 | 0.028**<br>[0.011] | 0.023**<br>[0.011] | 0.030**<br>[0.014] |
| Constant | 77.769***<br>[0.014] | 76.104***<br>[0.015] | 79.195***<br>[0.017] |
| N | 3,980,291 | 1,851,939 | 2,128,352 |

*Notes:* This table presents estimates of the effect of exposure to the de-worming campaign, first pooled then stratified by sex. Panel A presents coefficients from an alternative semi-parametric specification using the interaction between tertile of baseline hookworm prevalence and years of exposure, with coefficients relative to the lowest tertile. Panel B presents coefficients from a simplified pre-post framework, using those with less than 2 effective years of exposure as the reference group. Panel C most closely resembles the paper's primary specification, but uses a linear years of exposure before age 5 term, where the years of exposure are calculated as the birth year-1912, truncated at 0 on the low end and 5 on the high end. All empirical specifications reflect the primary specification using SEA fixed-effects that are allowed to vary by sex, and state-by-birth year fixed-effects. Data are drawn from the CenSoc BUNMD, weighted such that the distribution matches that from the Human Mortality Database. Hookworm prevalence is assigned at the SEA. Standard errors presented in brackets, clustered at the SEA level.

**Table A3:** Impacts of de-worming on lifespan, including additional controls.

|  |  | Longevity (years) |  |
| --- | --- | --- | --- |
|  | All | Males | Females |
| A. Add SEA rural quartile × birth-year controls | | | |
| SEA Prevalence × Effective Exposure Years | 0.038***<br>[0.013] | 0.033**<br>[0.015] | 0.040**<br>[0.018] |
| Constant | 77.769***<br>[0.012] | 76.102***<br>[0.016] | 79.195***<br>[0.016] |
| N | 3,980,291 | 1,851,939 | 2,128,352 |
| B. Add SEA rural, temperature, precipitation, and suitability for agriculture quartiles × birth year controls | | | |
| SEA Prevalence × Effective Exposure Years | 0.046***<br>[0.013] | 0.026<br>[0.017] | 0.055***<br>[0.017] |
| Constant | 77.768***<br>[0.013] | 76.112***<br>[0.018] | 79.189***<br>[0.015] |
| N | 3,960,391 | 1,842,320 | 2,118,071 |

*Notes:* This table presents estimates of the effect of exposure to the de-worming campaign, first pooled then stratified by sex. Empirical specifications follow the primary specification for mortality, using SEA fixed-effects that are allowed to vary by sex, and state-by-birth year fixed-effects. Additional characteristics of each SEA are divided into quartiles of the distribution and interacted with birth year, such that they absorb additional trends related to SEA characteristics that may be correlated with hookworm prevalence. Panel A presents coefficients including interactions between SEA rurality quartile calculated using the 1910 census (and thus available for all SEAs) and birth year. Panel B presents results that add to rurality a series of other indicators produced at the county level by Fiszbein, 2022 ((55)). I aggregate these to SEAs by taking averages weighted by area in an SEA. These characteristics include county mean temperature and precipitation, and GAO-FAEZ land suitability for cultivation. Lifespan data are drawn from the CenSoc BUNMD, weighted such that the distribution matches that from the Human Mortality Database. Hookworm prevalence is assigned at the SEA. Standard errors presented in brackets, clustered at the SEA level.

**Table A4:** Estimates of de-worming’s impact on mortality rates calculated in the US census.

|  |  | Survival |  |
| --- | --- | --- | --- |
|  | All | Males | Females |
| A. Primary specification |  |  |  |
| State Prevalence × Effective Exposure Years | 0.051*<br>[0.028] | 0.038<br>[0.029] | 0.064**<br>[0.026] |
| Constant | 0.770***<br>[0.008] | 0.738***<br>[0.008] | 0.804***<br>[0.007] |
| N | 341 | 341 | 341 |

*Notes:* This table presents estimates of the effect of exposure to the de-worming campaign, first pooled then stratified by sex. Observations only include RSC-treated states, and calculate the survival from early life (after infant mortality) until 1960 in the census. The empirical specification includes birth year and birth state fixed-effects. Standard errors presented in brackets, clustered at the state level.

**Table A5:**
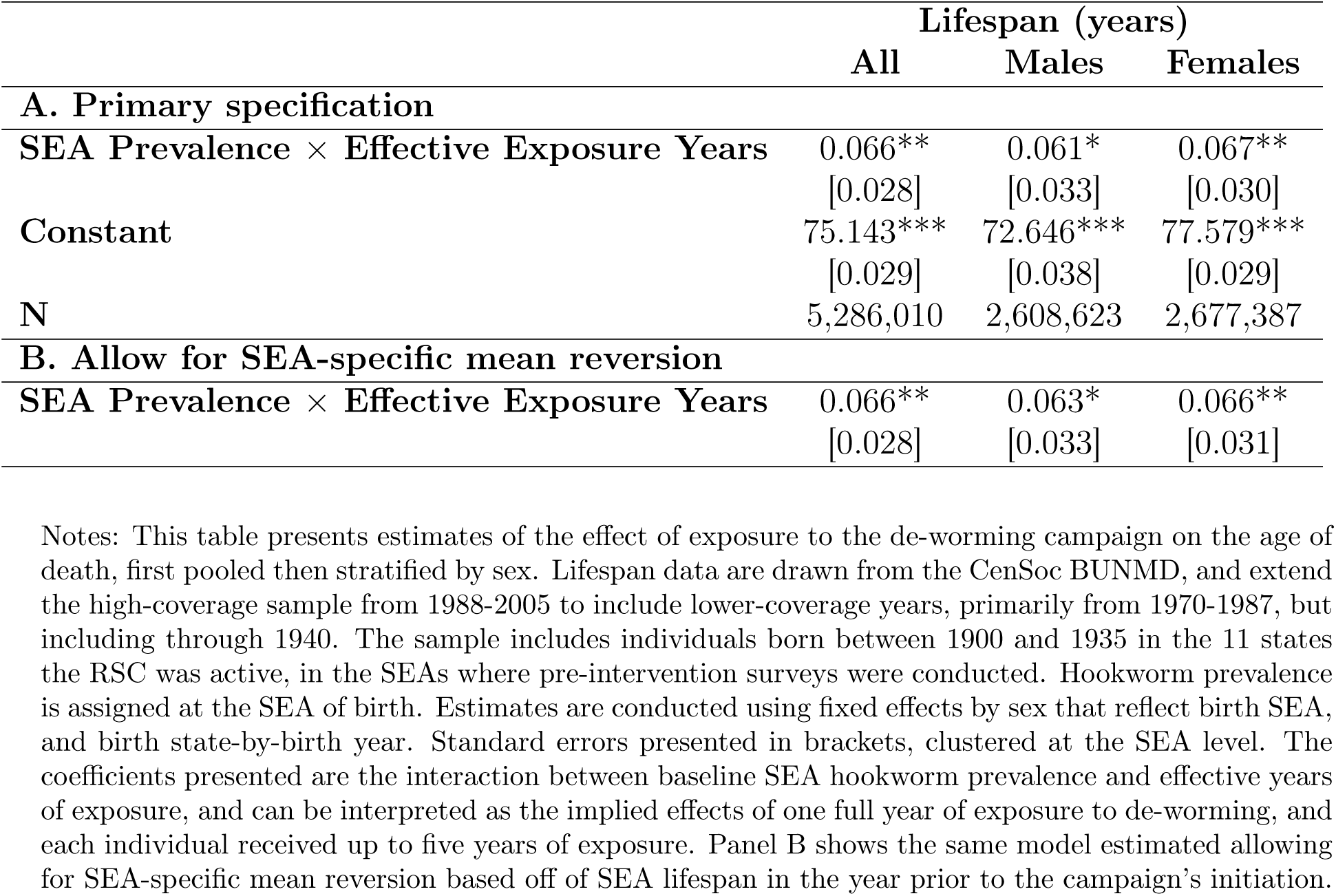
Estimated impacts of de-worming on lifespan, using extended sample data from the full Unified Numident, including low coverage years.

**Table A6:** Impacts of de-worming on BMI threshold probabilities.

| **A. Distributions of BMI** | BMI $\leq 20$ | | | BMI $\geq 25$ | | | BMI $\geq 30$ | | |
| --- | --- | --- | --- | --- | --- | --- | --- | --- | --- |
|  | All | M | F | All | M | F All | M | F |  |
| **Prevalence × Effective exposure years** | -0.054***<br>[0.020] | -0.068**<br>[0.031] | -0.038**<br>[0.016] | 0.078***<br>[0.018] | 0.091***<br>[0.024] | 0.052**<br>[0.022] | 0.032*<br>[0.018] | 0.014<br>[0.018] | 0.039<br>[0.025] |
| **N** | 17,450 | 8,162 | 9,288 | 17,450 | 8,162 | 9,288 | 17,450 | 8,162 | 9,288 |
| **Mean** | 0.144 | 0.142 | 0.146 | 0.476 | 0.492 | 0.464 | 0.152 | 0.119 | 0.181 |

Notes: This table presents estimates of the effect of exposure to the de-worming campaign, first pooled then stratified by sex. Pre-intervention hookworm prevalence is assigned by state of birth. Data are drawn from the NHANES I and NHANES II. Models include fixed effects for birth year and birth state, controls for sex, age, rural/urban, status, Black race, educational attainment, and low income level. The coefficients presented are the interaction between baseline state hookworm prevalence and effective years of exposure, and can be interpreted as the implied effects of one full year of exposure to de-worming, and each individual received up to five years of exposure. Outcomes reflect binary outcomes that reflect the impacts of de-worming on the probability of being at different points in the distribution, below 20, above 25, and above 30. Standard errors presented in brackets, clustered at the birth state level.

## Footnotes

1 Another potential concern is that mortality selection in childhood may impact adult outcomes - studies from diseases with high mortality rates may in part be picking up changes in mortality selection over time (*17*).

2 Several notable limitations have made testing the role of the hygiene hypothesis, and the role of helminths, challenging. Murine models have developed wild and wildling mice to test the hygiene hypothesis, finding limited support.(*24*). Models of helminth infection have implicated potentially important changes in microbiota(*25*). Other natural experiments from human populations in farming environments or birth order, while informative, are difficult contexts to isolate the impacts of a specific exposure (*26*, *27*). Studies of child birth order may also be contaminated by socio-economic impacts of birth order. (*28*)

3 Estimated shares of deaths covered closely follow counts in each birth cohort (Supplement figure A2).

4 Gender differences are consistent with women living longer than men, on average. Nonetheless, limited differences in the sample representation by sex suggest that endogenous selection on older-age mortality may not be particularly concerning in practice. Additionally, these data cover a large fraction of deaths generally, but do particularly well covering deaths around the RSC intervention.

5 Mean reversion is potentially a concern if areas had high hookworm and lower lifespan in the pre-period because of some transient shock. Table 2 panel B accounts for potential mean reversion by estimating the average lifespan for each birth SEA in 1912, and including in the primary specification a set of interactions between 1912 average lifespan in an SEA and effective exposure years.

6 Controls for birth year interacted with rurality, precipitation, temperature, and soil suitability capture potential trends related to characteristics that affect hookworm prevalence that may affect SEA lifespan trajectories over time. In practice, I produce quartiles of the distribution for each characteristic in the sample, and add birth year-by-quartile fixed effects to the priamry specification.

7 Increased standard errors also suggest increased variance of mortality in early life, compared with mortality in later life.

8 If adult socio-economic status and location is a potentially important route through which childhood de-worming affects adult morbidity and lifespan, these controls may be over-adjusting. However, empirical evidence of adult SES effects is small, and morbidity results are minimally-affected by inclusion of controls.

9 This study complements independent, simultaneous work by Noghanibehambari and Fletcher (*51*). Mortality findings are qualitatively similar, though several important differences are notable. I use a sample four times as large, and include both men and women, rather than just men. This is particularly important, as many effects are larger in women. Additionally, this study uniquely explores the long-term health impacts, and potential physiologic mechanisms by which de-worming may affect long-term mortality.

10 Kofoid notes that the burdens identified were the remaining light-intensity residual hookworm burden as hookworms in young soldiers died and they were not re-infected.

